# Modelling the recovery of elective waiting lists following COVID-19: scenario projections for England

**DOI:** 10.1101/2021.12.13.21267732

**Authors:** Nicholas C Howlett, Richard M Wood

## Abstract

**Background:** A significant indirect impact of COVID-19 has been the increasing elective waiting times observed in many countries. In England’s National Health Service, the waiting list has grown from 4.4 million in February 2020 to 5.7m by August 2021.

**Aims:** The objective of this study was to estimate the trajectory of future waiting list size and waiting times to December 2025.

**Methods:** A scenario analysis was performed using computer simulation and publicly available data as of November 2021. Future demand assumed a phased return of various proportions (0, 25, 50 and 75%) of the estimated 7.1 million referrals ‘missed’ during the pandemic. Future capacity assumed 90, 100 and 110% of that provided in the 12 months immediately before the pandemic.

**Results:** As a worst case, the waiting list would reach 13.6m (95% CI: 12.4m to 15.6m) by Autumn 2022, if 75% of missed referrals returned and only 90% of pre pandemic capacity could be achieved. Under this scenario, the proportion of patients waiting under 18 weeks would reduce from 67.6% in August 2021 to 42.2% (37.4% to 46.2%) with the number waiting over 52 weeks reaching 1.6m (0.8m to 3.1m) by Summer 2023. At this time, 29.0% (21.3% to 36.8%) of patients would be leaving the waiting list before treatment. Waiting lists would remain pressured under even the most optimistic of scenarios considered, with 18-week performance struggling to maintain 60% (against the 92% constitutional target).

**Conclusions:** This study reveals the long-term challenge for the NHS in recovering elective waiting lists as well as potential implications for patient outcomes and experience.

## 1. Introduction

The COVID-19 pandemic has exerted substantial pressure on healthcare services. While the direct impacts of the disease have mainly affected the acute emergency setting, the resulting reduction in hospital resources has constrained the capacity available for elective treatments. In order to manage the first wave of COVID-19 cases, many countries postponed all non-urgent procedures in public hospitals (Iacabucci et al, 2020; Babidge et al, 2021). Estimates suggest this contributed to over 28 million operations being cancelled or postponed globally (COVIDSurg Collaborative, 2020). This and subsequent pressures have led to the increase in waiting times that have been observed in many countries around the world (OECD, 2021).

Long waits are bad for both patients and healthcare providers. For patients, waiting time associates with worse health outcomes and can result in loss of independence, social isolation and depression (Hirvonen et al, 2009; Palagyi et al, 2016). Health inequalities, already increased during the pandemic (Bambra et al, 2020), risk further widening with those able to pay having the option to go private (Propper et al, 2020). For healthcare providers, additional and otherwise unnecessary costs may stem from the ongoing management of symptoms up until the point of definitive treatment (Wall et al, 2020). Costs may also be incurred for patients who, due to their waiting time, convert to the more disruptive and resource-intensive non-elective route – consider, for instance, the elevated fall risk of a delayed hip replacement or cataract patient (Harwood et al, 2005; Hill et al, 2016). Long waits also affect the economy more widely, given the reduced contribution to the workforce made by those with impaired functional mobility and worse health (Cole & Neumayer, 2006).

By Autumn 2021 (the time of writing), mass vaccination against COVID-19 has helped weaken the link between infection incidence and hospital bed occupancy (UK Cabinet Office, 2021; Yates, 2021). Yet while this has provided some stability for elective pathways, much uncertainty remains regarding the situation into 2022 and beyond. A substantial reduction in referrals has been observed in many countries during the pandemic (Morris et al, 2020; Zakeri et al, 2020; Tam et al, 2021): with societal restrictions now relaxed (given the vaccine), how many of these will now return? And although elective capacity has now mostly improved, will it be sufficient to effectively attack the backlog amassed in many healthcare systems? Especially if, while weakened, COVID-19 and associated infection control measures continue to restrict the hospital bed base beyond pre pandemic levels (Gogna et al, 2020; King & Hothi, 2021; UK Health Security Agency, 2021). There is also the matter of whether perennial recruitment issues in the health sector will limit the potential of additional funds pledged by many governments for accelerating elective recovery (UK Government, 2021).

These variables, essentially relating to elective care demand and capacity, can be flexed as part of scenario projections for future waiting list size and waiting times. Projections are important for a number of reasons. They help set realistic expectations for patients, enabling them to better plan their lives and make more informed decisions regarding alternative treatment options (such as conservative management or going private). They can be used by policy makers to more objectively assess the potential pros and cons of different resource allocation decisions, e.g. the diversion of extra healthcare funds to elective care may be warranted if projections reveal worse than considered waits. Also, they can be used to help estimate the additional capacity required to satisfy the aforementioned wait-driven healthcare demands, e.g. the number of new community physiotherapists required to manage ongoing symptoms or the extra emergency care resources needed for those converting to the non-elective route.

Against these possible benefits, there has been relatively little interest from the academic community in projecting waiting lists, especially during COVID-19 at the level needed to inform regional or national policy. Pre COVID-19, much of the effort involved bespoke projects at an individual hospital or specialty level, through use of operational research techniques such as queuing theory and computer simulation (George et al, 1983; Worthington, 1991; Azari-Rad et al, 2013; Dellaert et al, 2016). In very few models is the effect of long waits on patient health and behaviour captured and this is particularly important for modelling in the COVID-19 era, given the various pressures the pandemic has placed upon elective waiting lists.

The objective of this study is to estimate the trajectory of future waiting list size and waiting times in England’s National Health Service (NHS) to December 2025, under various scenarios considered plausible at the time of writing. The remainder of the paper is structured as follows. Section 2.1 details development and specification of a computer simulation to model the problem. Section 2.2 outlines the setting of the study and the range of scenarios considered. In Section 2.3, the calibration of the model is covered. Projections are presented in Section 3. Finally, a discussion on the strengths, limitations and practical implications is provided in Section 4.

## 2. Methods

### 2.1 Waiting list model

While the particular organisation of elective waiting lists will differ by country, specialty, and clinic (Hurst & Siciliani, 2003; Curtis et al, 2010), there are some general principles that can be used to guide model development. First, patients in need of treatment join the waiting list, typically following a GP (family doctor) referral. They are then treated according to their priority (based on clinical need), waiting time (elapsed time since referral), and complexity (in terms of healthcare activity required – some will require much more than others). Aggregate treatment rates are limited by available resources, e.g. diagnostic capacity, the number of consultants and surgeons, and operating theatre slots. Finally, some patients will leave the waiting list before treatment as waits increase.

Computer simulation is used to model this dynamical behaviour. The two key model inputs are the future number of referrals and capacity for treatment, both of which can be varied within a scenario analysis. Referrals are assumed Poisson-distributed around the inputted values, in order to appreciate referral rate variability and the independence of one referral to another. This is a common choice in modelling healthcare arrivals (Green, 2006). To capture patient complexity, referrals are partitioned on arrival to one of a number of classes, each of which awards a different number of points to the referral. The patients selected for treatment on each simulated day (bounded above by capacity for that day) are determined probabilistically with chances of selection equivalent to the number of points. This ensures that referrals of the highest-point classes, representing the least complex cases, have the greatest chance of being selected for treatment, and thus have the least waiting time. It also ensures that treatment order reflects waiting time, since it is more difficult for waiting patients to escape selection for treatment with each passing day.

As previously mentioned, patients may also leave the waiting list before treatment. This is known in queuing theory parlance as *reneging*. To address this, it is assumed that each waiting patient has a waiting time dependent probability of reneging, which is sampled on each simulated day. The probability is calculated using a sigmoidal function, whose value increases with waiting time. Specifically, the cumulative Weibull distribution function is used, with shape and scale parameter given by *k* and *λ* respectively. If the patient is determined to have reneged, they are removed from the waiting list. Within each simulated day, reneging occurs before treatment selection with new referrals being simulated last. This ensures that a patient cannot renege or be selected for treatment on the same day as their referral.

A single run of the simulation from the first to the last day in the considered projection period accounts for just one way in which events could pan out. To capture realistic variability and uncertainty, the random number seed used in the simulation loop is changed and another run is performed. This may mean a different number of arrivals on each day, a different selection of patients, and different patients reneging at different times. After a sufficient number of runs are performed, model outputs are combined to calculate the various projections of waiting list size, waiting times, and the number of patients reneging, for the scenario in question. Supplementary Material A contains the completed research checklist appropriate to this type of study (the STRESS guidelines, see Monks et al, 2019).

### 2.2 Study setting and scenarios

The above model is applied to the elective waiting list of England’s NHS as of August 2021 (the latest month of available data at the time of the study). At such time, elective waits had shown some signs of improvement although the backlog of patients awaiting treatment was still rising (Figure 1). Specifically, the 18-week referral-to-treatment (RTT) statistic, measuring the percentage of waiting patients waiting under 18 weeks, had improved from its 46.8% low in July 2020 and stabilised at approximately 68%. The RTT metric is the principal barometer of elective performance in the NHS and is assessed against a 92% constitutional target (UK Government, 2012). It is increasingly more instructive to consider the numbers waiting over 52 weeks, which increased 270-fold from February 2020 (before the pandemic) to its peak in March 2021. All data was obtained from a publicly available central NHS source (NHS England, 2021).

**Figure 1.**
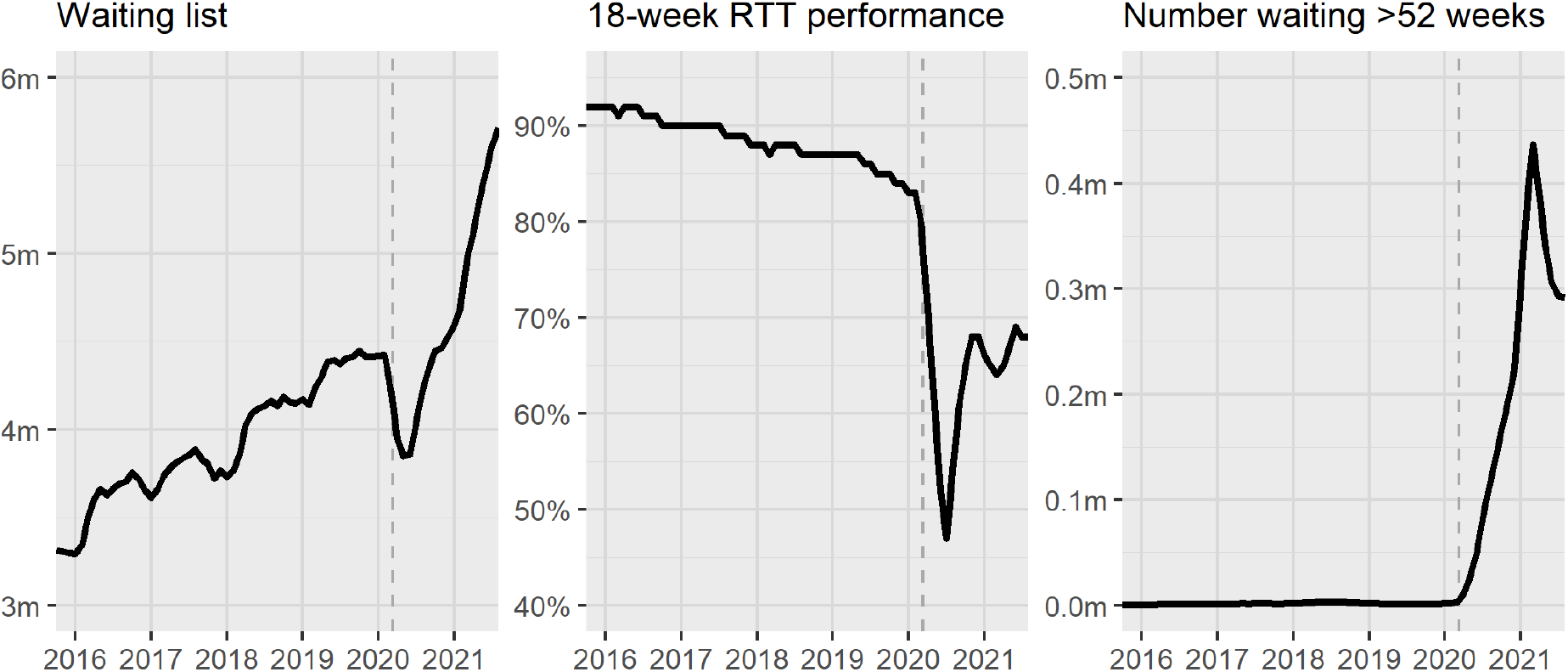
Elective care performance in England’s NHS to August 2021. The dashed grey vertical lines represent the declaration of a pandemic by the World Health Organisation on 11 March 2020.

The scenarios examined through the model (Table 1) are defined over the period from September 2021 to December 2025, and relate to the two previously mentioned ‘unknowns’ at the time of the study: the proportion of missed referrals returning and future capacity for treatment (Section 1). While monthly referrals had returned to pre pandemic levels by August 2021, an estimated 7.1 million had been ‘missed’ since March 2020 (NHS England, 2021). In our analysis, we consider that 0%, 25%, 50% and 75% of these will return within the 12 months from September 2021 (normally distributed, µ=182 days, σ^2^=55, and truncated between 0 and 365 days), on top of a ‘baseline’ referral rate equivalent to the mean from March 2019 to February 2020 (i.e. the last full 12 months before the pandemic). Also considered is an increasing referral growth rate based upon the rate observed in the five years pre pandemic, which is linear (p < 0.001) and increases by approximately 50,000 each year (this represents 2.8% annual growth in the latest pre pandemic year). Relative to total elective treatments from March 2019 to February 2020, we consider future capacity at 90%, 100% and 110% for the period until December 2025 (phased in over the first six months). The lower limit reflects constrained elective capacity assuming sustained COVID-19 demand, while the upper limit accounts for success in government plans to increase elective capacity (UK Government, 2021). No clear cyclical behaviour could be detected in monthly elective treatments and so no seasonality is assumed for capacity.

**Table 1.** Scenarios examined within this study. Note that approximately 7.1 million elective referrals were ‘missed’ during the pandemic from March 2020 to August 2021 and that 16.6m treatments were performed in the 12 months to February 2020 (the last full year before COVID-19 was declared a pandemic).

| Scenario | Proportion of referrals ‘missed’ during the pandemic to return in 12 months from September 2021 | Elective capacity relative to the number of treatments performed in 12 months to February 2020 |
| --- | --- | --- |
| 1 | 0% | 110% |
| 2 | 0% | 100% |
| 3 | 0% | 90% |
| 4 | 25% | 110% |
| 5 | 25% | 100% |
| 6 | 25% | 90% |
| 7 | 50% | 110% |
| 8 | 50% | 100% |
| 9 | 50% | 90% |
| 10 | 75% | 110% |
| 11 | 75% | 100% |
| 12 | 75% | 90% |

### 2.3 Model calibration

For parsimony, two classes are assumed for representing patient complexity, each of which must be assigned a probability of selection and a number of points (Section 2.1). Given that assignment to Class A and B are complementary, it is necessary to define only one probability parameter (e.g. if Class A probability is 0.8 then Class B probability is 0.2). Given also that it is the relative difference in points that informs selection of patients for treatment, it is only necessary to define the number of points awarded to one class (here the other is set equal to one).

These two model parameters relating to the complexity classes, alongside the two relating to the reneging function (*k* and *λ*), were estimated through fitting to the latest six months data for waiting list size, 18-week performance, and numbers waiting over 52 weeks (Figure 1). Accuracy was determined through Mean Absolute Percentage Error (MAPE), with larger weights applied to more recent data (according to an exponential decay function with a one-month half-life). Modelling upon the actual referrals and capacity over the six-month period (NHS England, 2021), the accuracy of one million distinct parameter sets was computed. For each of the one million simulations, MAPE was calculated from the mean of 100 performed runs. All computational processes used R version 4.1.1 on a Microsoft Windows machine (i9-9980XE CPU, 64GB RAM).

Of the one million parameter sets, the top 1000 (by MAPE) were selected for use in obtaining the scenario projections (Section 3). This limited overreliance on one or a small number of parameterisations, which could otherwise promote undue confidence in model outputs. To reasonably address such uncertainty, projections included the mean of the 1000 trajectories and 95% confidence bands based on the 2.5% and 97.5% quantiles. Supplementary Material B contains a comparison of model projections and actual values over the six-month calibration period. This supports goodness of fit for waiting list size and 18-week performance, with all actual values contained within the modelled confidence bands. This also holds for the most recent four months of the number waiting over 52 weeks, although there is some larger discrepancy for the first two months. Parameter estimates, calculated from the top 1000 selected parameter sets, give a 0.90 mean (95% interval: 0.86 to 0.92) for the probability of a referral being assigned to complexity Class A with a 110 mean (10 to 200) points awarded. One interpretation of this larger and less complex class is a lack of need for significant surgical procedure, which is supported by a similarly large proportion of elective treatments being carried out without hospital admission (NHS England, 2021). The reneging parameter estimates give a 2.8 mean (2.4 to 3.0) for *k* and a 2654 mean (2025 to 3000) for *λ*. The correspondence between all model parameters is detailed in Supplementary Material C.

## 3. Results

Assuming no long-term trend growth in referrals, the waiting list would peak at 13.6m (95% CI: 12.4m to 15.6m) in Autumn 2022 (Figures 2 and 3) if 75% of referrals missed during the pandemic returned in the 12 months from September 2021 and only 90% of pre pandemic elective capacity could be provided (Scenario 12, Table 1). If 110% capacity can be achieved, then peak waiting list size would be reduced by approximately two million (Scenario 10). If none of the missed referrals returned then, under the most optimistic scenario of those considered, the waiting list would reduce over the longer term, reaching 5.2m (3.0m to 6.7m) by December 2025 (Scenario 1). In general, results show that regardless of the number of missed referrals that return, the waiting list will over time stabilise to levels determined by capacity (Figures 2 and 4). This is due to higher rates of reneging from the waiting list in the shorter term when the waiting list – and moreover waiting times – are larger. This is most prominent for Scenario 12, where the proportion of patients reneging increases three-fold to peak at 29.0% (21.3% to 36.8%) in Spring 2023. Note that the estimated reneging function, inferred through model calibration (Section 2.3), can be found in Supplementary Material D.

**Figure 2.**
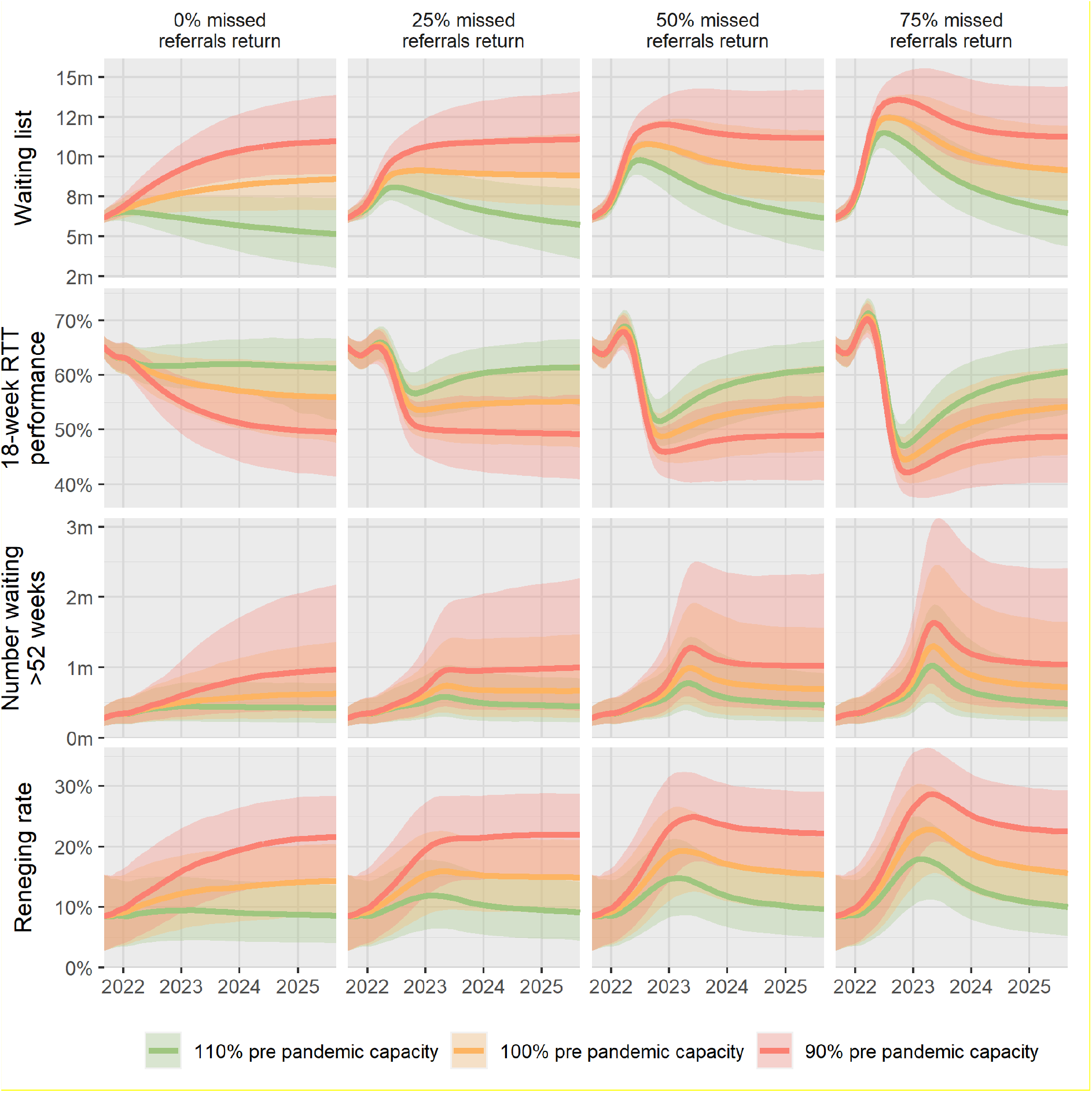
Model projections, accounting for the pre pandemic referral rate (March 2019 to February 2020) with no trend growth and various proportions of the estimated 7.1 million referrals ‘missed’ during the pandemic returning in the 12 months from September 2021.

**Figure 3.**
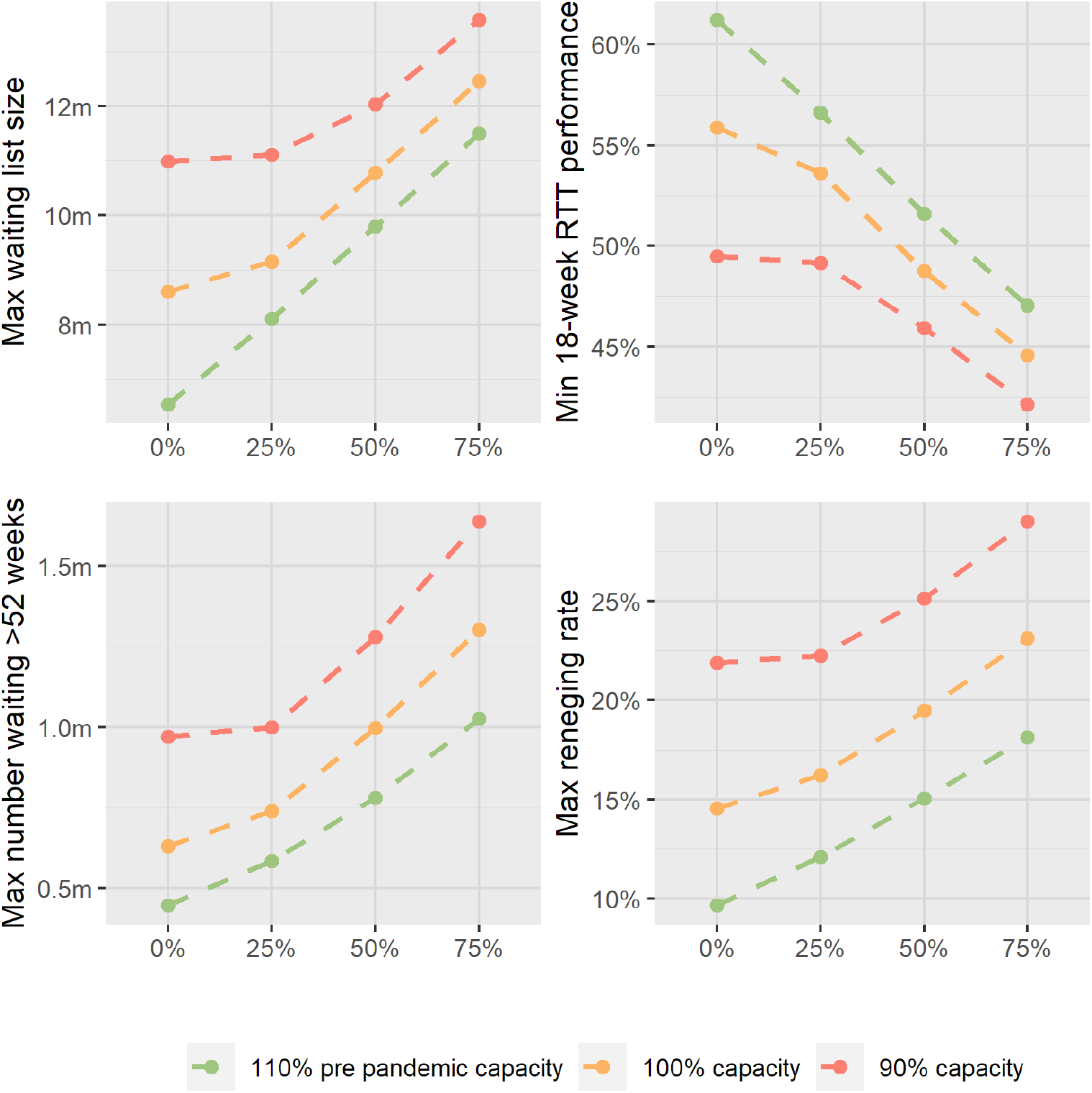
Peak values of model projections from September 2021 to December 2025, accounting for the pre pandemic referral rate (March 2019 to February 2020) with no trend growth and various proportions of the estimated 7.1 million referrals ‘missed’ during the pandemic returning in the 12 months from September 2021.

**Figure 4.**
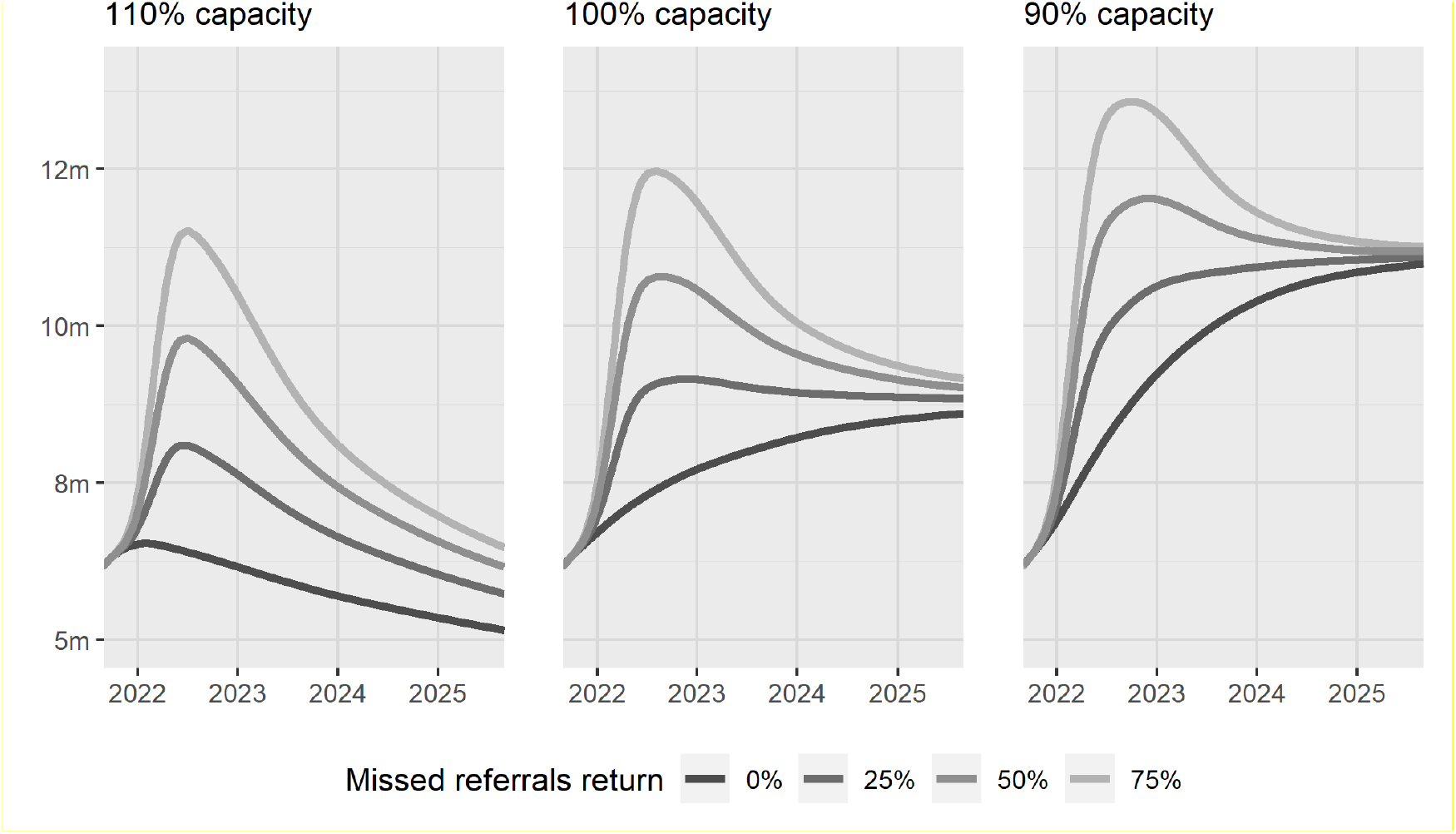
Long term convergence of waiting list projections across the considered capacities, accounting for the pre pandemic referral rate (March 2019 to February 2020) with no trend growth and various proportions of the estimated 7.1 million referrals ‘missed’ during the pandemic returning in the 12 months from September 2021.

If 75% of missed referrals returned, then waiting time performance could fall below 50% when measured by the 18-week RTT metric (Scenarios 10-12). This compares to the 46.8% low recoded in July 2020 (Figure 1). Over the longer term, above 60% performance can be achieved provided 110% capacity can be sustained. Indeed, above 60% performance could be maintained throughout the period should no missed referrals return. Note that the brief spikes in 18-week performance in Spring 2022, most noticeable for scenarios involving a greater return of missed referrals, are an artefact of the 18-week RTT equation, given the flood of new referrals which temporarily increase the numerator (number of patients waiting < 18 weeks). Although the 18-week metric represents the constitutional target, it is increasingly more instructive to look at numbers waiting over 52 weeks. As a worst case, this would peak at 1.6m (0.8m to 3.1m) in Summer 2023 (Scenario 12). If 110% capacity is provided, numbers can be limited to within 0.5m over the longer term (c.f. 436,127 recorded in February 2020).

Accounting for referral growth at the rate observed in the five years pre pandemic, results indicate a much greater challenge in restoring elective performance (Figure 5). Even under the most optimistic of scenarios considered, modelling suggests that capacity would be insufficient to prevent a sustained increase in waiting list size and deterioration in patient waiting times over the longer term.

**Figure 5.**
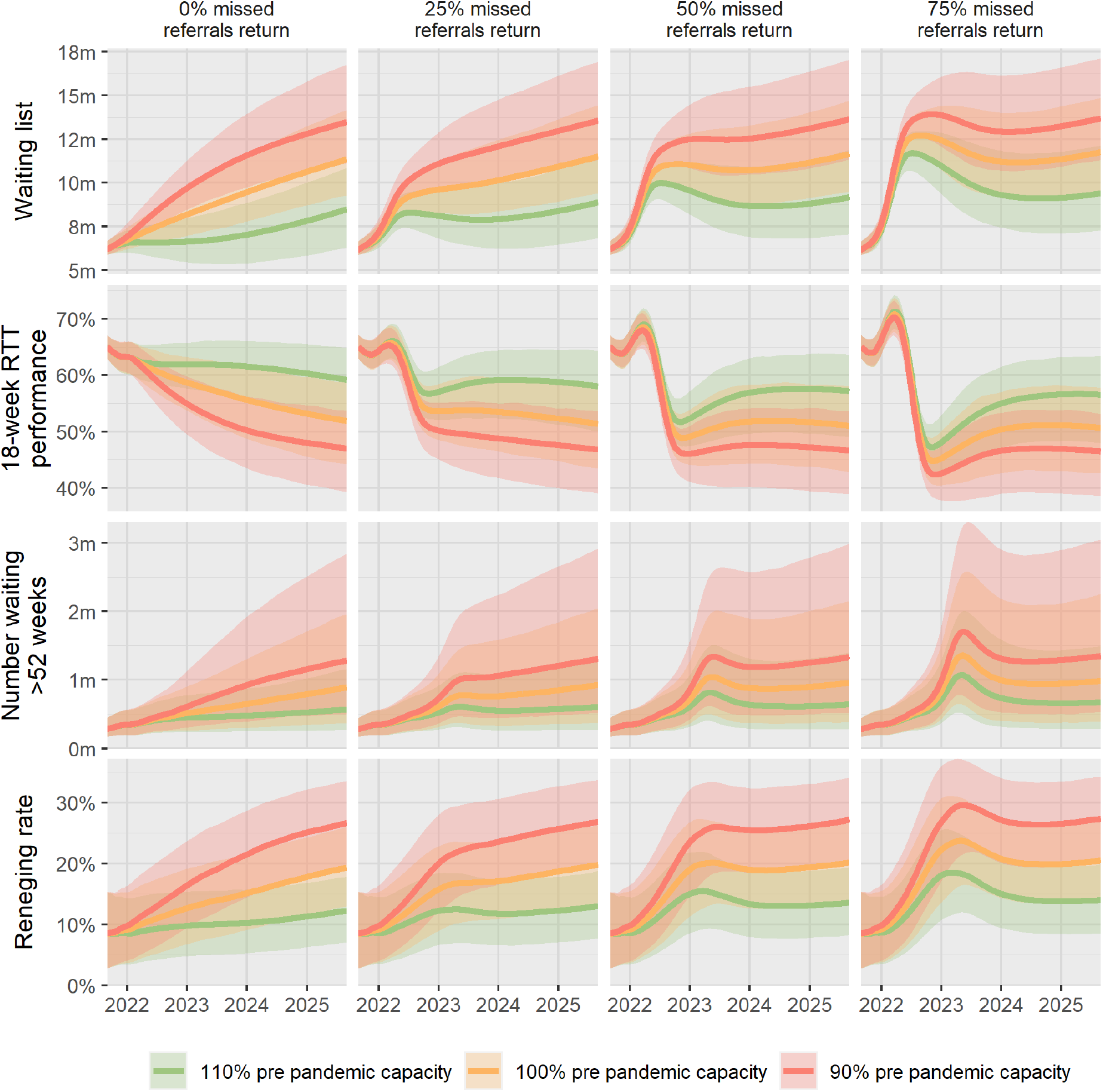
Model projections, accounting for referral growth at the rate observed in the five years pre pandemic (from March 2015 to February 2020) and various proportions of the estimated 7.1 million referrals ‘missed’ during the pandemic returning in the 12 months from September 2021.

## 4. Discussion

Despite the significant indirect impact of COVID-19 on elective performance, and the attendant media interest and public concern, there has been a deficit of applied modelling to project the future state of waiting lists and what this could mean for patients and healthcare services. Indeed, at the time of this study, no other modelling work could be found in the peer-reviewed literature concerning the recovery of elective waiting lists in England.

The results of this study reveal the long-term challenge for the NHS in recovering elective waiting lists following the early impacts of COVID-19. Even without accounting for a long-term trend growth in referrals, the projections presented here will raise numerous questions for healthcare systems (Figure 2). How will ongoing symptoms be managed over such long waits, and at what cost? What additional resources will be consumed by patients converting, as a consequence of waiting time, to the non-elective route? How could health inequalities be reduced if those with the means to do so decide to go private? And to what extent will longer term impairments to population health affect macro-economic indicators such as unemployment and growth? The need to resolve these questions will be greater should referrals grow according to the longer-term trend evident before the pandemic (Figure 5).

Increasing capacity is a possible solution to address waiting list growth and improve waiting times. In October 2021, the UK Treasury pledged a further £6b to address the elective backlog in England, on top of £8b already earmarked over three years (Anandaciva, 2021). We calculate, using an assumed £2,933 average elective pathway cost (Wood, 2021), that this £14b would buy extra capacity to treat a total 4.8m patients, equivalent to 1.6m annually. This amount is approximately equivalent to the additional requirement for scenarios involving 110% of pre pandemic capacity, given the 16.6m treatments performed in the 12 months prior to the pandemic (NHS England, 2021). However, our modelling suggests that more investment would be required to improve long term 18-week performance much beyond the 60% level. As well as financial budget, attracting and retaining a suitable workforce is another challenge that must be met to increase capacity (Olive, 2020). Other options include improving productivity and, of course, reducing demand, potentially through greater promotion of conservative management approaches (Macdonald et al, 2020).

Turning to limitations, although a satisfactory model fit to historical data has been possible for waiting list size and 18-week performance, there is a less convincing approximation to numbers waiting over 52 weeks (Section 2.3). Further work may be used to explore whether greater accuracy could be achieved with the use of additional complexity classes, beyond the two considered here. It should also be acknowledged that while patient complexity is captured, the current model does not appreciate referral priority and its influence on treatment order. Future investigators may wish to explore the extent to which incorporation of this may improve model accuracy. However, given the various ways that different specialties and clinics prioritise treatment for waiting patients (Hurst & Siciliani, 2003; Curtis et al, 2010), it is likely to be difficult to capture priority within models applied at a national, specialty-wide level.

## Data Availability

Model input data is available online at https://www.england.nhs.uk/statistics/statistical-work-areas/rtt-waiting-times/. Model output data is available upon reasonable request to the authors.

