## supplementary material for "Modelling the recovery of elective waiting lists following COVID-19: scenario projections for England"

### Supplementary Material A

Strengthening the Reporting of Empirical Simulation Studies (STRESS)
Discrete-event simulation guidelines STRESS-DES

| Section/Subsection | Item | Recommendation | Location in article |
|---|---|---|---|
| **1. Objectives** | | | |
| Purpose of the model | 1.1 | Explain the background and objectives for the model. | Section 1 |
| Model Outputs | 1.2 | Define all quantitative performance measures that are reported, using equations where necessary. Specify how and when they are calculated during the model run along with how any measures of error such as confidence intervals are calculated. | Section 2.1 |
| Experimentation Aims | 1.3 | If the model has been used for experimentation, state the objectives that it was used to investigate.<br><br>a.) Scenario based analysis – Provide a name and description for each scenario, providing a rationale for the choice of scenarios and ensure that item 2.3 (below) is completed.<br><br>b.) Design of experiments – Provide details of the overall design of the experiments with reference to performance measures and their parameters (provide further details in *data* below).<br><br>c.) Simulation Optimisation – (if appropriate) Provide full details of what is to be optimised, the parameters that were included and the algorithm(s) that was be used. Where possible provide a citation of the algorithm(s). | Section 2.2 |
| **2. Logic** | | | |
| Base model overview diagram | 2.1 | Describe the base model using appropriate diagrams and description. This could include one or more process flow, activity cycle or equivalent diagrams sufficient to describe the model to readers. Avoid complicated diagrams in the main text. The goal is to describe the breadth and depth of the model with respect to the system being studied. | Section 2.1 |
| Base model logic | 2.2 | Give details of the base model logic. Give additional model logic details sufficient to communicate to the reader how the model works. | Section 2.1 |

| | | | | |
|---|---|---|---|---|
| Scenario logic | 2.3 | Give details of the logical difference between the base case model and scenarios (if any). This could be incorporated as text or where differences are substantial could be incorporated in the same manner as 2.2. | | Section 2.2 |
| Algorithms | 2.4 | Provide further detail on any algorithms in the model that (for example) mimic complex or manual processes in the real world (i.e. scheduling of arrivals/appointments/operations/maintenance, operation of a conveyor system, machine breakdowns, etc.). Sufficient detail should be included (or referred to in other published work) for the algorithms to be reproducible. Pseudo-code may be used to describe an algorithm. | | Section 2.1 |
| Components | 2.5 | 2.5.1 Entities | Give details of all entities within the simulation including a description of their role in the model and a description of all their attributes. | Section 2.1 |
| | | 2.5.2 Activities | Describe the activities that entities engage in within the model. Provide details of entity routing into and out of the activity. | Section 2.1 |
| | | 2.5.3 Resources | List all the resources included within the model and which activities make use of them. | Section 2.1 |
| | | 2.5.4 Queues | Give details of the assumed queuing discipline used in the model (e.g. First in First Out, Last in First Out, prioritisation, etc.). Where one or more queues have a different discipline from the rest, provide a list of queues, indicating the queuing discipline used for each. If reneging, balking or jockeying occur, etc., provide details of the rules. Detail any delays or capacity constraints on the queues. | Section 2.1 |
| | | 2.5.5 Entry/Exit Points | Give details of the model boundaries i.e. all arrival and exit points of entities. Detail the arrival mechanism (e.g. 'thinning' to mimic a non-homogenous Poisson process or balking) | Section 2.1 |

| | | | |
|---|---|---|---|
| 3. **Data** | | | |
| Data sources | 3.1 | List and detail all data sources. Sources may include:<br><br>• Interviews with stakeholders,<br>• Samples of routinely collected data,<br>• Prospectively collected samples for the purpose of the simulation study,<br>• Public domain data published in either academic or organisational literature. Provide, where possible, the link and DOI to the data or reference to published literature.<br><br>All data source descriptions should include details of the sample size, sample date ranges and use within the study. | Section 2.2 |
| Pre-processing | 3.2 | Provide details of any data manipulation that has taken place before its use in the simulation, e.g. interpolation to account for missing data or the removal of outliers. | Section 2.2 |
| Input parameters | 3.3 | List all input variables in the model. Provide a description of their use and include parameter values. For stochastic inputs provide details of any continuous, discrete or empirical distributions used along with all associated parameters. Give details of all time dependent parameters and correlation.<br><br>Clearly state:<br><br>• Base case data<br>• Data use in experimentation, where different from the base case.<br>• Where optimisation or design of experiments has been used, state the range of values that parameters can take.<br><br>Where theoretical distributions are used, state how these were selected and prioritised above other candidate distributions. | Section 2.2 |
| Assumptions | 3.4 | Where data or knowledge of the real system is unavailable what assumptions are included in the model? This might include parameter values, distributions or routing logic within the model. | N/A |
| 4. **Experimentation** | | | |

| | | | |
|---|---|---|---|
| Initialisation | 4.1 | Report if the system modelled is terminating or non-terminating. State if a warm-up period has been used, its length and the analysis method used to select it. For terminating systems state the stopping condition.<br><br>State what if any initial model conditions have been included, e.g., pre-loaded queues and activities. Report whether initialisation of these variables is deterministic or stochastic. | Section 2.3 |
| Run length | 4.2 | Detail the run length of the simulation model and time units. | Section 2.3 |
| Estimation approach | 4.3 | State the method used to account for the stochasticity: For example, two common methods are multiple replications or batch means. Where multiple replications have been used, state the number of replications and for batch means, indicate the batch length and whether the batch means procedure is standard, spaced or overlapping. For both procedures provide a justification for the methods used and the number of replications/size of batches. | Section 2.3 |
| 5. **Implementation** | | | |
| Software or programming language | 5.1 | State the operating system and version and build number.<br><br>State the name, version and build number of commercial or open source DES software that the model is implemented in.<br><br>State the name and version of general-purpose programming languages used (e.g. Python 3.5).<br><br>Where frameworks and libraries have been used provide all details including version numbers. | Section 2.3 |
| Random sampling | 5.2 | State the algorithm used to generate random samples in the software/programming language used e.g. Mersenne Twister.<br><br>If common random numbers are used, state how seeds (or random number streams) are distributed among sampling processes. | Section 2.1 |
| Model execution | 5.3 | State the event processing mechanism used e.g. three phase, event, activity, process interaction.<br><br>*Note that in some commercial software the event processing mechanism may not be published. In these cases authors should adhere to item 5.1 software recommendations.* | Section 2.1 |

| | | | |
|---|---|---|---|
| | | State all priority rules included if entities/activities compete for resources.<br><br>If the model is parallel, distributed and/or use grid or cloud computing, etc., state and preferably reference the technology used. For parallel and distributed simulations the time management algorithms used. If the HLA is used then state the version of the standard, which run-time infrastructure (and version), and any supporting documents (FOMs, etc.) | |
| System Specification | 5.4 | State the model run time and specification of hardware used. This is particularly important for large scale models that require substantial computing power. For parallel, distributed and/or use grid or cloud computing, etc. state the details of all systems used in the implementation (processors, network, etc.) | Section 2.3 |
| **6. Code Access** | | | |
| Computer Model Sharing Statement | 6.1 | Describe how someone could obtain the model described in the paper, the simulation software and any other associated software (or hardware) needed to reproduce the results. Provide, where possible, the link and DOIs to these. | Contact corresponding author |

**Supplementary Material B**

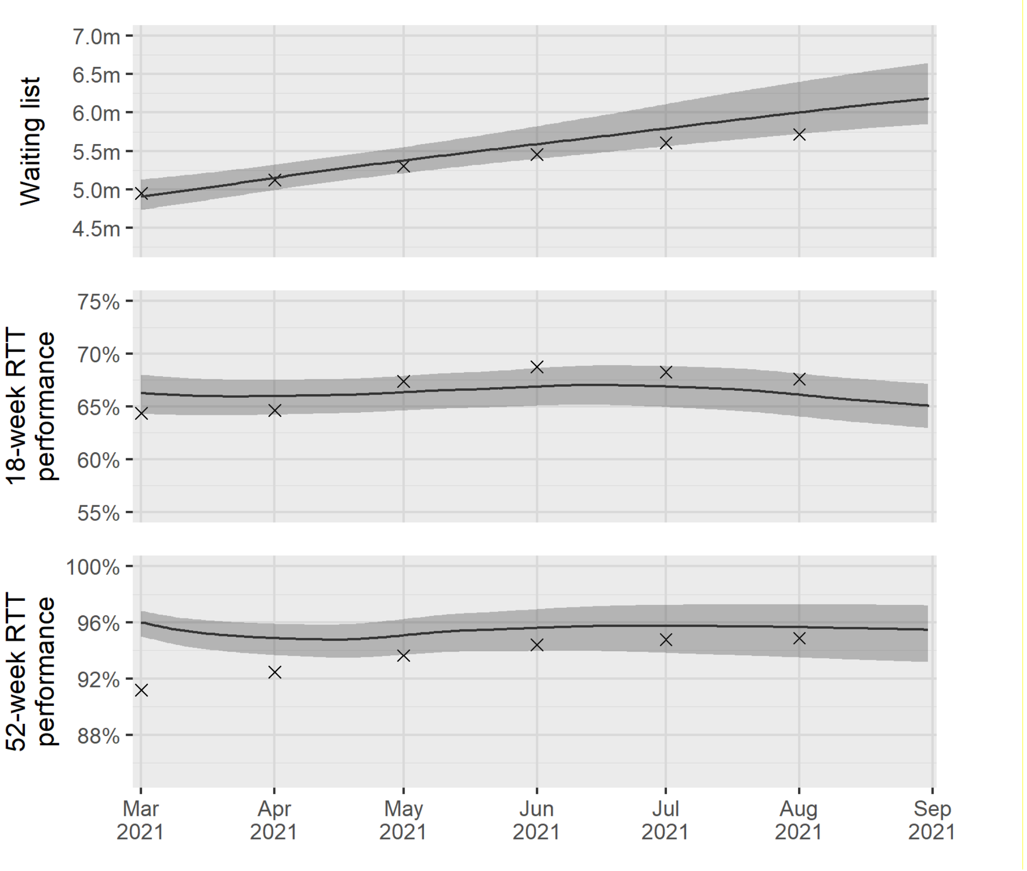


**Figure SM.B.** Comparison of actual values (crosses) and model projections (mean and 95% confidence bands) as determined from the top 1000 parameter sets, over the six-month calibration period.

**Supplementary Material C**

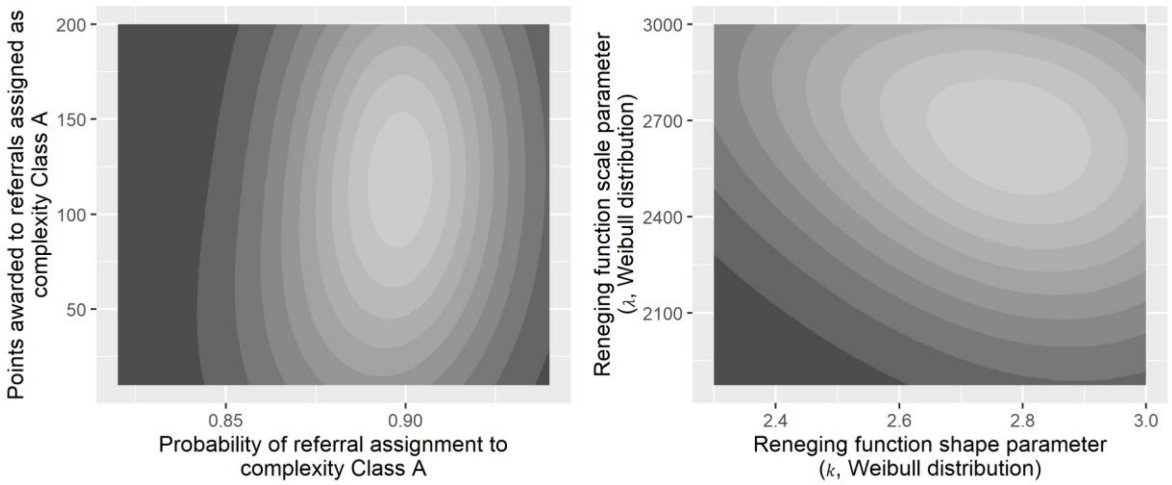


**Figure SM.C.** Correspondence between the parameters relating to the two complexity classes and the reneging function, as determined from the top 1000 parameter sets selected for use in obtaining scenario projections. Lighter shading represents areas of the parameter space with greater model accuracy (as determined by MAPE against historical actual data).

**Supplementary Material D**

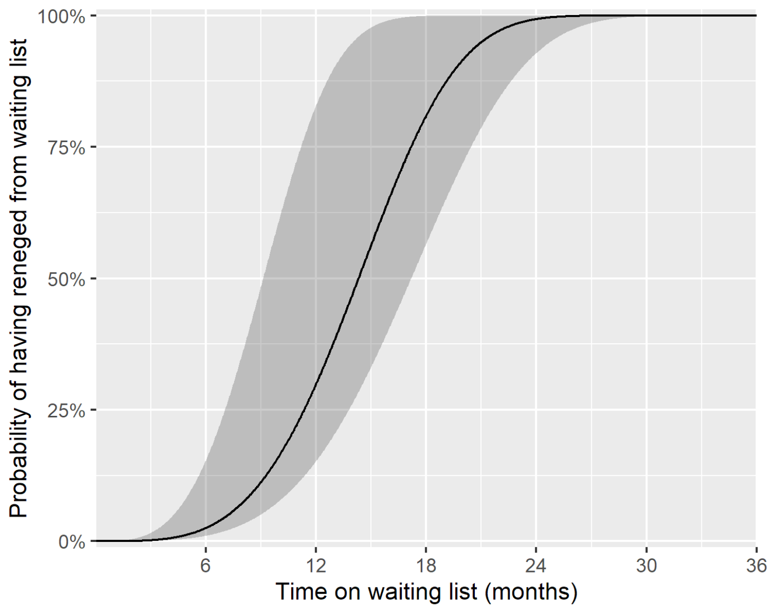


**Figure SM.D.** Inferred probability of a patient having reneged from the waiting list before treatment given time spent on the waiting list. Results, including the mean (solid black line) and 95% interval (shaded area), are obtained from the Weibull cumulative distribution function calibrated through estimates of $k$ and $\lambda$ from the top 1000 parameter sets. Results are conditional on the patient having not been selected for treatment.
